# Exposure–response relationships for personal exposure to fine particulate matter (PM_2·5_), carbon monoxide, and black carbon and birthweight: Results from the multi-country Household Air Pollution Intervention Network (HAPIN) trial

**DOI:** 10.1101/2022.08.06.22278373

**Authors:** Kalpana Balakrishnan, Kyle Steenland, Thomas Clasen, Howard Chang, Michael Johnson, Ajay Pillarisetti, Wenlu Ye, Luke P. Naeher, Anaite Diaz-Artiga, John P. McCracken, Lisa M. Thompson, Ghislaine Rosa, Miles A. Kirby, Gurusamy Thangavel, Sankar Sambandam, Krishnendu Mukhopadhyay, Naveen Puttaswamy, Vigneswari Aravindalochanan, Sarada Garg, Florien Ndagijimana, Stella Hartinger, Lindsay UnderHill, Katherine A Kearns, Devan Campbell, Jacob Kremer, Lance Waller, Shirin Jabbarzadeh, Jiantong Wang, Yunyun Chen, Joshua Rosenthal, Ashlinn Quinn, Aris T. Papageorghiou, Usha Ramakrishnan, Penelope P. Howards, William Checkley, Jennifer L. Peel, HAPIN Investigators

## Abstract

**Background:** Household air pollution (HAP) from solid fuel use is associated with adverse birth outcomes, but data on exposure-response relationships are limited. We examined associations between HAP exposures and birthweight in rural Guatemala, India, Peru, and Rwanda during the Household Air Pollution Intervention Network (HAPIN) trial.

**Methods:** We recruited 3200 pregnant women between 9 and <20 weeks of gestation. Women randomized to the intervention arm received a liquified petroleum gas (LPG) stove and fuel during pregnancy, while control arm women continued using biomass. We measured 24-hr personal exposures to particulate matter (PM_2·5_), carbon monoxide (CO), and black carbon (BC) once pre-intervention (baseline), twice post-intervention, and birthweight within 24 hours of birth. We examined the relationship between the average prenatal exposure and birthweight/weight-for-gestational age z-scores using multivariate-regression models.

**Findings:** Results showed an inter-quartile increase in average prenatal exposure to PM_2.5_ (74·5 μg/m^3^) and BC (7·3 μg/m^3^) was associated with a 14·8 (95% confidence interval [CI]: -28·7g, - 0·8g) and 21·9g (95% CI: -37·3g, -6·1g) reduction in birthweight and reduced weight-for-gestational age z-scores of -0·03 (95% CI: -0.06, 0·00) and -0·05 (95%CI: -0·08, -0·01) standard deviations, respectively. We found no associations for birthweight or weight-for-gestational age z-scores with CO exposures.

**Interpretation:** Results provide support for continuing efforts to reduce HAP exposure alongside other drivers of low birthweight in low- and middle-income countries.

**Funding:** The study is registered with ClinicalTrials.gov (NCT02944682) and funded by the U.S. National Institutes of Health (1UM1HL134590) in collaboration with the Bill & Melinda Gates Foundation (OPP1131279).

## Introduction

Household air pollution (HAP) exposures from the use of solid cooking fuels such as wood, coal, charcoal, dung, and agricultural residues are a leading risk factor for ill-health in low- and middle-income countries (LMICs), accounting for an estimated 2·3 million premature deaths annually and 91·5 million disability-adjusted life years.^1^ Systematic reviews have summarized the evidence for an association between HAP exposure and adverse health effects, including child pneumonia, chronic obstructive lung disease, lung cancer, and cataracts.^2^ Relatively few studies or reviews have focused on adverse perinatal outcomes including low birthweight.^3–6^

LMICs bear a disproportionate share of low birthweight (LBW, defined as <2500 g regardless of gestational age), accounting for nearly 91% of the global burden.^7^ The etiology of LBW is complex, and despite ongoing efforts to address known risk factors such as maternal malnutrition, malaria, and smoking,^8^ progress has been slow towards the ambitious global nutrition target of a 30% reduction of LBW by 2025.^7^ As nearly 3·8 billon people worldwide rely on solid fuels,^9^ a strengthened understanding of the relationship between HAP and LBW would be extremely valuable for prioritizing efforts to decrease HAP exposures during pregnancy to improve birth outcomes.

Most previous studies that examine the association between HAP exposures and LBW have used categorical indicators of exposure based on primary fuel use, with only a handful reporting quantitative exposure-response (E-R) relationships for particulate matter with an aerodynamic diameter ≤ 2·5 micrometers (PM_2·5_)^10,11^ or carbon monoxide (CO).^12–14^ These E-R studies report significant associations between prenatal PM_2·5_ and/or CO exposures and LBW, but also report many limitations: small sample sizes, an inability to measure multiple pollutants, and the use of single personal exposure measures during pregnancy and/or longitudinal kitchen area measurements as proxies of longer-term personal exposure. Recent randomized control trials (RCTs) of HAP interventions in Nepal,^15^ Nigeria,^12^ and Ghana^16^ have reported null effects from intention-to-treat analyses for impacts on birthweight, but E-R analyses within these studies have been limited.^13^ To our knowledge, no studies have examined E-R relationships between prenatal black carbon (BC) exposures and birthweight.

We present results from E-R analyses performed as part of the Household Air Pollution Intervention Network (HAPIN) RCT, an efficacy study of a free prenatal liquefied petroleum gas (LPG) stove and fuel intervention conducted across four LMICs (Guatemala, India, Peru, Rwanda) with repeated personal exposure measurements. We hypothesized that higher pregnancy period PM_2·5_, BC, and CO exposures would result in lower birthweight among infants born to mothers enrolled in the HAPIN trial in each of – and across – the four countries.

## Methods

### Study participants and settings

Participants were pregnant women enrolled in the HAPIN trial, details of which have been published previously^17–19^ and are summarized in the trial registration (ClinicalTrials.gov Identifier NCT02944682). The specific study areas in each country (Jalapa Municipality, Guatemala; Villupuram and Nagapatinam districts of Tamil Nadu, India; Department of Puno, Peru; and Eastern Province, Rwanda) were selected based on high prevalence of cooking with biomass, low background ambient PM_2·5_ concentrations, and acceptable field feasibility as assessed during an 18-month period of planning and formative research.^20,21^ Between March 2018 and February 2020, we recruited a total of 3200 (800 per country) non-smoking, pregnant women who were between 18 and ≤35 years of age, between 9 and ≤ 20 weeks of gestation (determined via ultrasound), and who used biomass as a primary fuel. In accordance with the trial protocol, half of the participants in each country were randomized to an intervention arm that received a liquefied petroleum gas (LPG) stove and a continuous supply of LPG fuel following enrollment and throughout their pregnancy, while the balance served as controls and continued to rely chiefly on solid biomass for cooking.

### Personal exposure monitoring during pregnancy

Prenatal personal exposure monitoring protocols and results have been described previously.^19,22^ Briefly, at each study site, pregnant women participated in three 24-hr personal exposure assessments, once at baseline (between 9 and <20 weeks of gestation) and twice after randomization into the control or intervention arms (between 24-32 weeks and 32-36 weeks of gestation, respectively). During each session, women wore customized vests or aprons fitted so that instrumentation was situated close to their breathing zone. PM_2·5_ monitoring was performed using the Enhanced Children’s MicroPEM™ (ECM) (RTI International), which collects (a) gravimetric samples on pre-weighed 15mm Teflon filters (MTL,USA) utilizing a 2·5 micron impactor at a flow rate of 0·3 liters per minute and (b) real-time nephelometric data.^23^ BC was estimated post-sampling on the ECM filters using the SootScan® Model OT-21 Optical Transmissometer (Magee Scientific, USA). CO monitoring was performed using the Lascar EL-CO-USB-300 DataLogger (Lascar Electronics, USA). Participants were instructed to always wear the vest or apron during the 24-hr measurement period, except when sleeping, bathing, or when conducting other activities during which the equipment could not be safely worn. During these times, they were instructed to keep the vest or apron nearby. Additionally, data were collected on sociodemographic and household characteristics and activity patterns that may influence exposure.

Procedures for assuring data quality, weighing filters, and estimating missing gravimetric data based on nephelometry have been described previously.^22^ Briefly, gravimetric data quality assurance involved a combination of threshold values for flow rates, inlet pressure, and sampling duration, as well as visual inspection of damaged filters by weighing room technicians. In cases where nephelometric but not gravimetric data were available, PM_2·5_ exposure was estimated based on nephelometric data, using an instrument-specific regression coefficient for the association between nephelometric and gravimetric data for that specific ECM instrument as described previously.^22^ CO quality assurance protocols included calibrations with zero air and span gas and a visual inspection system similar to what was applied in the GRAPHS trial in Ghana.^24^

For E-R analyses, gestational exposures were defined for the intervention group as the average of the pre- and post-intervention exposures, weighted by the amount of gestational time spent in each period. The pre-intervention period exposure was estimated using the baseline measurement, while the post-intervention exposure was estimated using one or both personal measurements performed after intervention. This allowed for exposure changes resulting from the introduction of the intervention to be weighted according to the length of time participants had the intervention during gestation. An unweighted average of the baseline and other available (1-2) gestational period measurements was used for controls, as they continued using biomass as the primary cooking fuel throughout gestation.

### Birthweight Outcome Measurements

Following a standard protocol, birthweight was measured within 24 hours of birth by a trained field worker or nurse using a Seca 334 mobile digital baby scale. Newborns were weighed naked to the nearest 10 g and duplicate measurements were recorded on tablet-based REDCap forms. If the first two measured birthweights differed by more than 10 g, a third measurement was taken. The average of the measurements was used in the data analysis. Infants were typically assessed at health facilities where they were born. Each scale was calibrated weekly in the field offices before deployment using standard 5-lb and 10-lb weights; scales not within ±2·5% of the standard weight were re-calibrated. In cases in which we were unable to reach the child during the prescribed 24-hr window—due mainly to COVID restrictions or critically ill infants admitted to ICUs or referral hospitals—we used measurements provided by the facility, if available, but conducted sensitivity analyses to compare results.

As gestational age is a potential mediator in the causal pathway between HAP exposure and birthweight, we did not adjust for it in the E-R models; had we done so, its inclusion would not allow estimation of the total effect of exposure.^25^ However, we additionally estimated z-scores for weight adjusted by gestational age defined using INTERGROWTH tables (intergrowth21.tghn.org) as a secondary analysis. These weight-for-gestational age z-scores were derived by subtracting off the standard INTERGROWTH sex-specific weight for a given gestational age and dividing by the INTERGROWTH standard deviation of that weight. Measurements were considered invalid if the gestational age at birth was greater than 300 days or if the birth weight-for-gestational age z-score did not fall between -6 and 5.

### Exposure-Response Modeling

The statistical analysis plan was agreed upon in advance and published with the trial registration prior to unblinding. Analyses were independently replicated by a second member of the study team. E-R analyses were modeled separately for each pollutant (PM_2·5_, BC, and CO) and birthweight/birthweight-for-gestational age z-score.

Covariate selection for models was guided by a directed acyclic graph (DAG) (Figure S3). A minimal set of potential confounders or strong risk factors (e.g., infant sex) were identified in systematic reviews of birthweight,^3,6^ and from previous studies of HAP and birthweight.^10–12,24,26^ We used 10% change-in-estimate (CIE) methods as outlined in Greenland (1989)^27^ to evaluate and determine covariates included in the model. Final models included the following covariates: mother’s age (categorical: <20/20-24/25-29/30-35), nulliparity (categorical: yes/no), diet diversity (categorical: low/median/high), food insecurity (categorical: food secure/mild/moderate), baseline BMI (continuous), mother’s education (categorical), child gender (categorical), baseline hemoglobin (continuous), and second-hand smoke (categorical: yes/no). We also included a variable for ten geographical randomization strata (one in Rwanda, one in Guatemala, two in India, six in Peru). Eighteen and twenty-six subjects were missing BMI and hemoglobin, respectively, and we created a category of missing for these variables so that they were not excluded from the analysis.

For both birthweight and z-scores, we first fitted linear models with different exposure metrics (i.e., linear, log linear). We then evaluated nonlinear categorical (quartile modes), as well as quadratic, 2-piece linear and restricted cubic spline model with three knots^28^ models, and assessed model fit using Akaike’s Information Criterion (AIC). The knots for the 2-piece spline were chosen based on AIC (using quartile cutpoints initially and then narrowing down), while knots for restricted cubic splines were placed at the 5^th^, 50^th^, and 75^th^ percentiles of exposure. We also used thin plate smoothing splines via generalized additive models, with penalization determined by generalized cross-validation score, using R package *mgcv*. We also examined effect modification by country, as well as by infant sex, via interaction terms between our exposure metrics and these variables.

### Ethics/registration/funding

The study protocol was reviewed and approved by institutional review boards (IRBs) or Ethics Committees at Emory University (00089799), Johns Hopkins University (00007403), Sri Ramachandra Institute of Higher Education and Research (IEC-N1/16/JUL/54/49) and the Indian Council of Medical Research – Health Ministry Screening Committee (5/8/4-30/(Env)/Indo-US/2016-NCD-I), Universidad del Valle de Guatemala (146-08-2016/11-2016) and Guatemalan Ministry of Health National Ethics Committee (11-2016), A.B. PRISMA, the London School of Hygiene and Tropical Medicine (11664-5) and the Rwandan National Ethics Committee (No·357/RNEC/2018), and Washington University in St. Louis (201611159). The study was funded by the U.S. National Institutes of Health (1UM1HL134590) in collaboration with the Bill & Melinda Gates Foundation (OPP1131279). The funding sources were not involved in study design, collection, analysis, and interpretation of data, or decisions to submit the paper for publication.

## Results

### Participant Characteristics

While 3200 women were enrolled in the study, 5 enrollees were determined to be ineligible after randomization and exited the study. After accounting for miscarriages, stillbirth, and withdrawals, the 3195 pregnancies yielded 3060 live births. Of these, 3018 had valid birthweights (others had birthweights measured outside the 24-hr window or the study team was unable to obtain any birthweight measurement, see CONSORT diagram, Figure S1). Sixteen additional births were excluded on account of a gestational age >300 days; weight-for-gestational age z-scores are unavailable in the INTERGROWTH database beyond 300 days of gestation. 3002 subjects were thus eligible for inclusion in E-R analyses. These were further restricted by the availability of exposure data for each of the three pollutants of interest (Table 1).

**Table 1.**
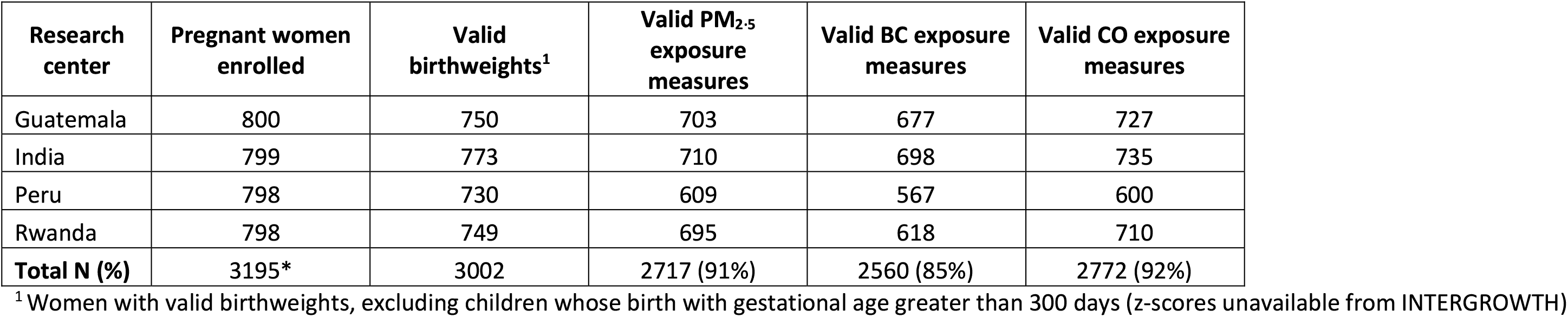
Summary of number of observations used in the exposure-response analysis

Participant characteristics are summarized in Table 2. The average age was 27·6 years, with 38% of participants reporting nulliparity. The average (SD) gestational age at enrollment was 15·3 (± 3·1) weeks. Only 33% women reported acquiring secondary or higher levels of education. India had the lowest BMI, hemoglobin, and diet diversity scores, while Peru had the highest. India had the highest proportion of smokers in the household. Mobile phone ownership was uniformly high across countries.

**Table 2.** Trial-wide and country-specific maternal characteristics

| **Maternal characteristics<sup>1</sup>** | **Guatemala** | **India** | **Peru** | **Rwanda** | **Total** |
| --- | --- | --- | --- | --- | --- |
|  | **N=750** | **N=773** | **N=730** | **N=749** | **N=3002** |
| Age (years), N (%) |  |  |  |  |  |
| <20 | 115 (15) | 122 (15) | 93 (12) | 46 (6) | 376 (12) |
| 20 – 24 | 303 (40) | 373 (48) | 261 (35) | 187 (25) | 1124 (37) |
| 25-29 | 221 (29) | 223 (28) | 232 (31) | 280 (37) | 956 (31) |
| 30-35 | 111 (15) | 55 (7) | 144 (19) | 236 (31) | 546 (18) |
| Gestational age at recruitment (weeks), mean (SD) | 14.3 (3) | 16 (3) | 15.7 (3.3) | 15.4 (2.8) | 15.3 (3.1) |
| Nulliparous, N (%) |  |  |  |  |  |
| Yes | 213 (28) | 442 (57) | 278 (38) | 218 (29) | 1151 (38) |
| No | 537 (71) | 337 (42) | 448 (61) | 529 (70) | 1845 (61) |
| Highest level of education completed, N (%) |  |  |  |  |  |
| No formal education / some primary school | 358 (47) | 275 (35) | 32 (4) | 316 (42) | 981 (32) |
| Primary school / some secondary school | 298 (38) | 219 (28) | 224 (30) | 299 (40) | 1034 (35) |
| Secondary / Vocational /Some University | 100 (13) | 279 (36) | 474 (65) | 134 (18) | 987 (33) |
| Height (cm), mean (SD) | 148 (5.3) | 151 (5.6) | 152 (4.5) | 156 (5.8) | 152 (6.2) |
| Body mass index (kg/m<sup>2</sup>) mean (SD) | 23.7 (3.3) | 19.7 (3.1) | 26 (3.5) | 23.4 (3.4) | 23.2 (4.1) |
| Hemoglobin (gm/dl), mean (SD) | 12.7 (1.04) | 10.3 (1.2) | 14 (1.2) | 12.4 (1.5) | 12.4 (1.9) |
| Minimum dietary diversity, Category (score) N (%) |  |  |  |  |  |
| Low (<4) | 514 (68) | 600 (77) | 73 (10) | 505 (67) | 1692 (56) |
| Medium (4-5) | 206 (27) | 149 (20) | 403 (55) | 208 (27) | 966 (32) |
| High (>5) | 30 (4) | 24 (3) | 254 (34) | 35 (4) | 343 (11) |
| Household food insecurity, Category (score), N (%) |  |  |  |  |  |
| Food secure | 415 (56) | 628 (81) | 378 (52) | 276 (37) | 1697 (57) |
| Mild (1, 2, 3) | 238 (32) | 108 (14) | 251 (34) | 212 (29) | 809 (27) |
| Moderate (4, 5, 6) / Severe (7, 8) | 88 (11) | 33 (4) | 91 (12) | 243 (33) | 455 (15) |
| Number of people sleeping in house, mean (SD) | 5·1 (2·6) | 3·7 (1·5) | 4·5 (1·7) | 3·4 (1·4) | 4·3 (2) |
| Someone in the household smokes, N (%) |  |  |  |  |  |
| Yes | 39 (5) | 244 (31) | 7 (1) | 28 (4) | 318 (11) |
| No | 711 (94) | 529 (68) | 722 (99) | 719 (96) | 2681 (89) |
| Owns household assets, N (%) |  |  |  |  |  |
| Color Television | 344 (45) | 577 (74) | 470 (64) | 98 (13) | 1489 (49) |
| Radio | 283 (37) | 105 (13) | 540 (73) | 420 (56) | 1348 (45) |
| Mobile phone | 687 (91) | 635 (82) | 699 (95) | 594 (79) | 2815 (87) |
| Bicycle | 94 (12) | 120 (15) | 278 (38) | 229 (30) | 721 (24) |
| Bank account | 186 (24) | 695 (89) | 172 (23) | 221 (29) | 1274 (42) |

<sup>1</sup> Descriptive statistics summary based on 3002 pregnant women included in the final analyses, which includes women with live births, valid birthweights, and gestational age at birth < 300 days

### PM_2·5_, BC, and CO exposures

We obtained 2717, 2560, and 2772 valid 24-hr prenatal personal PM _2·5_, BC, and CO exposure measurements, respectively. Mean (SD) weighted exposures during pregnancy were 92·2 (83·9) μg/m^3^ for PM_2·5_, 10·0 (7·4) μg/m^3^ for BC, and 2.0 (2·9) ppm for CO (Table S6). PM_2·5_ and BC exposures were highly correlated (Spearman’s *ρ* = 0·79), but correlations between exposure to PM_2·5_ and CO (Spearman’s *ρ* = 0·34) as well as BC and CO (Spearman’s *ρ* = 0·39) were relatively weak. The intervention resulted in marked reduction in exposure. Post-intervention mean personal PM_2·5_ was 24.0 μg/m^3^ in the intervention arm and 70.7μg/m^3^ in the control arm. Similar reductions of exposure were seen for BC and CO.

Exposure distributions are depicted in Figure 1 and described in Supplemental Table 6. Details on exposure settings and additional sociodemographic characteristics are reported elsewhere.^22^ Missing exposure data was largely due to equipment failure and was likely to be missing at random (MAR); for more details see Johnson et al. (2021).^22^

**Figure 1.**
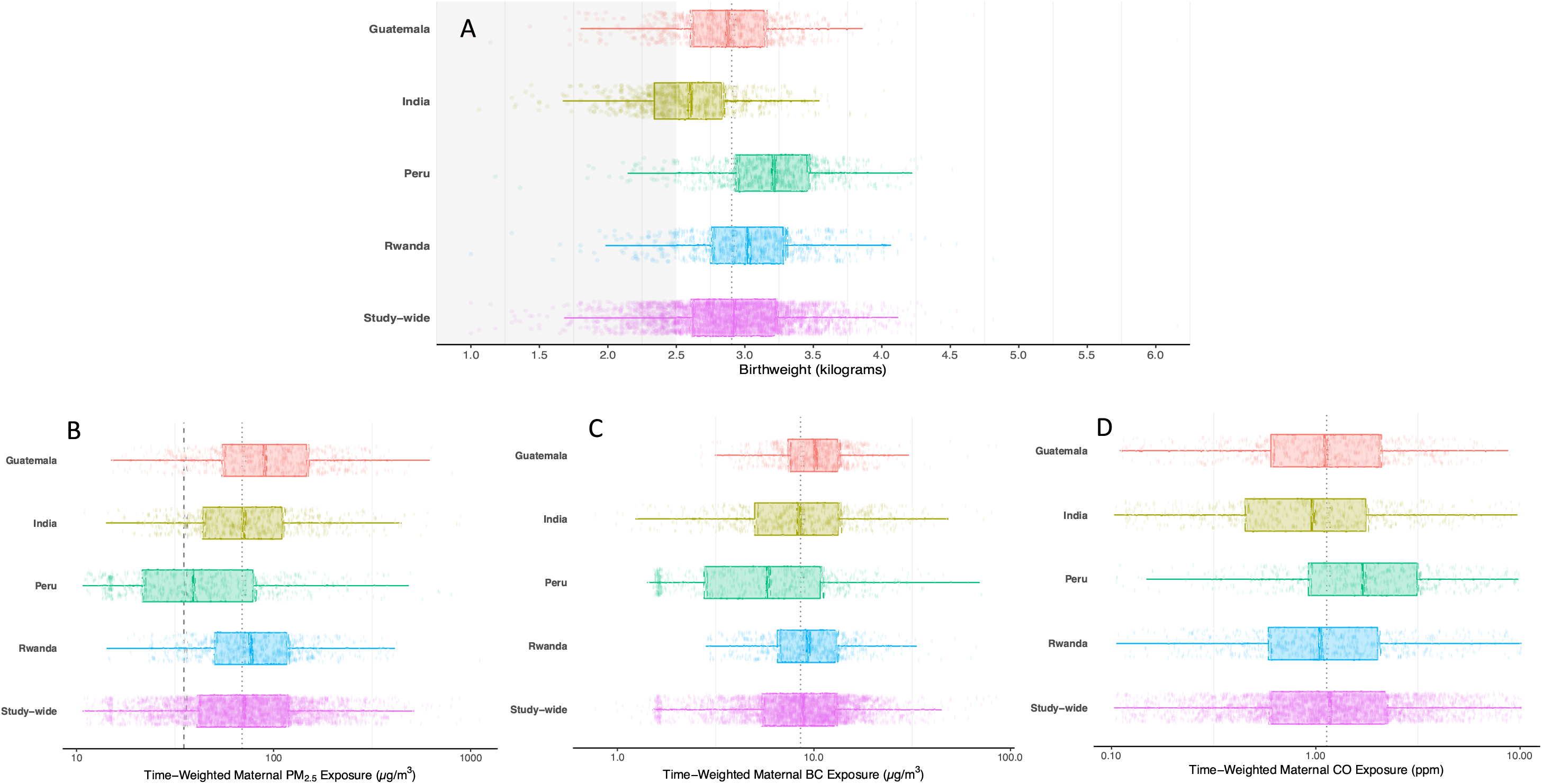
Distribution of (A) birthweight and time-weighted (B) PM_2·5_, (C) BC, and (D) CO. Corresponding numeric data are in Supplemental Table 6. Results are presented separately for each study site and in combination for the entire trial. Dots are individual datapoints. X-axes are log transformed. Thick solid lines inside the box are medians. The lower and upper hinges (i.e., the ends of the box) correspond to the 25^th^ and 75^th^ percentiles. The whiskers (i.e., the lines beyond the box) extend from the hinge to 1·5 * IQR. The panel-wide dotted vertical lines are study-wide medians. In panel (A), the shaded area indicates low birthweight (< 2500 g). In Panel (B), the dashed line is the WHO Interim Target 1 annual guideline value of 35 μg/m3.

### Birthweight

The mean (SD) birthweight of live born infants was 2909 (471) g with mean gestational age at delivery of 39·3 (1·5) weeks; 5·3% of births were classified as preterm (163/3002) and 17·7% as LBW (Figure 1). Mean (SD) birthweight was 2921 g (474·3 g) in the intervention arm and 2898 g (467·9 g) in the control arm, a difference of 19·6 g (95% CI: -10·1 g, 49·2 g).

### Exposure-response analyses

Results for linear and log-linear models for birthweight and for weight-for-gestational age z-score are shown in Table 3 and Table 4, respectively, for each of the three measured pollutants. Quartile models are presented in Tables S1 and S2. In linear models, an inter-quartile increase in gestational exposure for PM_2·5_ (74·5 μg/m^3^) and BC (7·3 μg/m^3^) was associated with a change in birthweight of -14.8 g (95% CI: -28·7 g, -0·8 g] and -21·9 g (95% CI: -37·7 g, -6·1 g], respectively (Table 3). For weight-for-gestational age z-scores, the same exposure increases were associated with a decrease of 0·03 (95% CI: -0·06, 0·00) and 0·05 (95%CI: -0·08, -0·01) standard deviations, respectively (Table 4). No associations were apparent between CO exposures and birthweight in the linear models or between any of the measured pollutants and LBW prevalence. Quartile analyses (Tables S1 and S2) showed that the decrease in birthweight and z-scores were not monotonic for PM_2.5_, while decreases were monotonic for z-scores but not birthweight for BC.

**Table 3.**
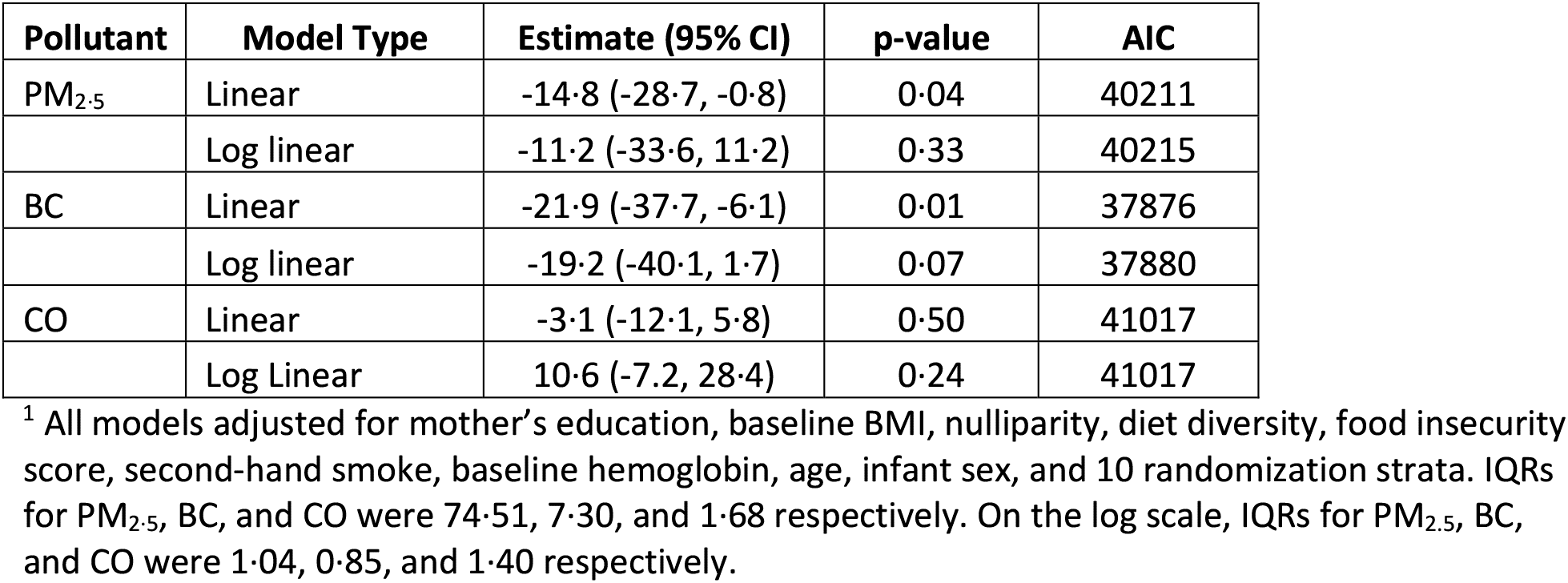
Change in birthweight for an IQR increase in PM_2·5_, BC, and CO^1^

**Table 4.** Change in weight-for-gestational age z-scores with an IQR increase in PM_2·5_, BC, and CO^1^

| Pollutant | Model | Estimate (95% CI) | p-value | AIC |
| --- | --- | --- | --- | --- |
| PM<sub>2.5</sub> | Linear | -0.03 (-0.06, 0.00) | 0.04 | 7021 |
|  | Log linear | -0.04 (0.09, 0.01) | 0.10 | 7023 |
| BC | linear | -0.05 (-0.08, -0.01) | 0.01 | 6591 |
|  | Log linear | -0.06 (-0.10, -0.01) | 0.02 | 6593 |
| CO | linear | -0.003 (-0.023, 0.017) | 0.78 | 7215 |
|  | Log linear | 0.02 (-0.02, 0.06) | 0.24 | 7214 |

<sup>1</sup> All models adjusted for mother's education, baseline BMI, nulliparity, diet diversity, food insecurity score, second-hand smoke, baseline hemoglobin, age, infant sex and 10 randomization strata. IQRs for PM<sub>2.5</sub>, BC, and CO were 74.51, 7.30, and 1.68 respectively. On the log scale, IQRs for PM<sub>2.5</sub>, BC, and CO were 1.04, 0.85, and 1.40 respectively.

Evaluation of different models indicated that the linear fit presented above was appropriate to model the relationships between the birthweight outcomes and BC. For PM_2.5_, however, a quadratic (non-linear) fit was better suited to the birthweight outcome (Table S3 and Figure S2), with a positive linear coefficient (0·2325) and a negative quadratic coefficient (-0·009), indicating an initial increase in birthweight with higher PM_2·5_ followed by a subsequent decrease at the higher exposures. Both categorical and cubic spline models supported this relationship (Figure S2). Linear models fit best for BC for both birthweight and z-scores, as well as PM_2.5_ and z-scores. Smoothed E-R curves for PM_2·5_/BC and birthweight and weight-for-gestational age z-scores can be seen in Figure 2 and Figure 3. Trends for full term births (95% of births) were similar to trends for all births (Table S4). No statistically significant interactions (at the 0·05 level) were observed with infant sex, but female births showed a larger effect than male births for birthweight, and for z-scores (Table S5). Trends were reasonably consistent across countries for the association between PM_2.5_ and BC with both birthweight and z-scores (Tables S7 and S8). We also ran separate models for our three exposure measurements during gestation, i.e. for baseline, mid-point, and end of gestation measurements (these corresponding roughly to early 2^nd^ trimester, end of 2^nd^ trimester, and end of 3^rd^ trimester). These models, for both birthweight and z-score, showed no pattern whereby early or later exposures had stronger effects on the outcome (Table S9). Indeed all time-specific E-R coefficients were weaker than those coefficients using average exposure. This might occur because single measurements involve more measurement error than average exposure across gestation, biasing results to the null.

**Figure 2.**
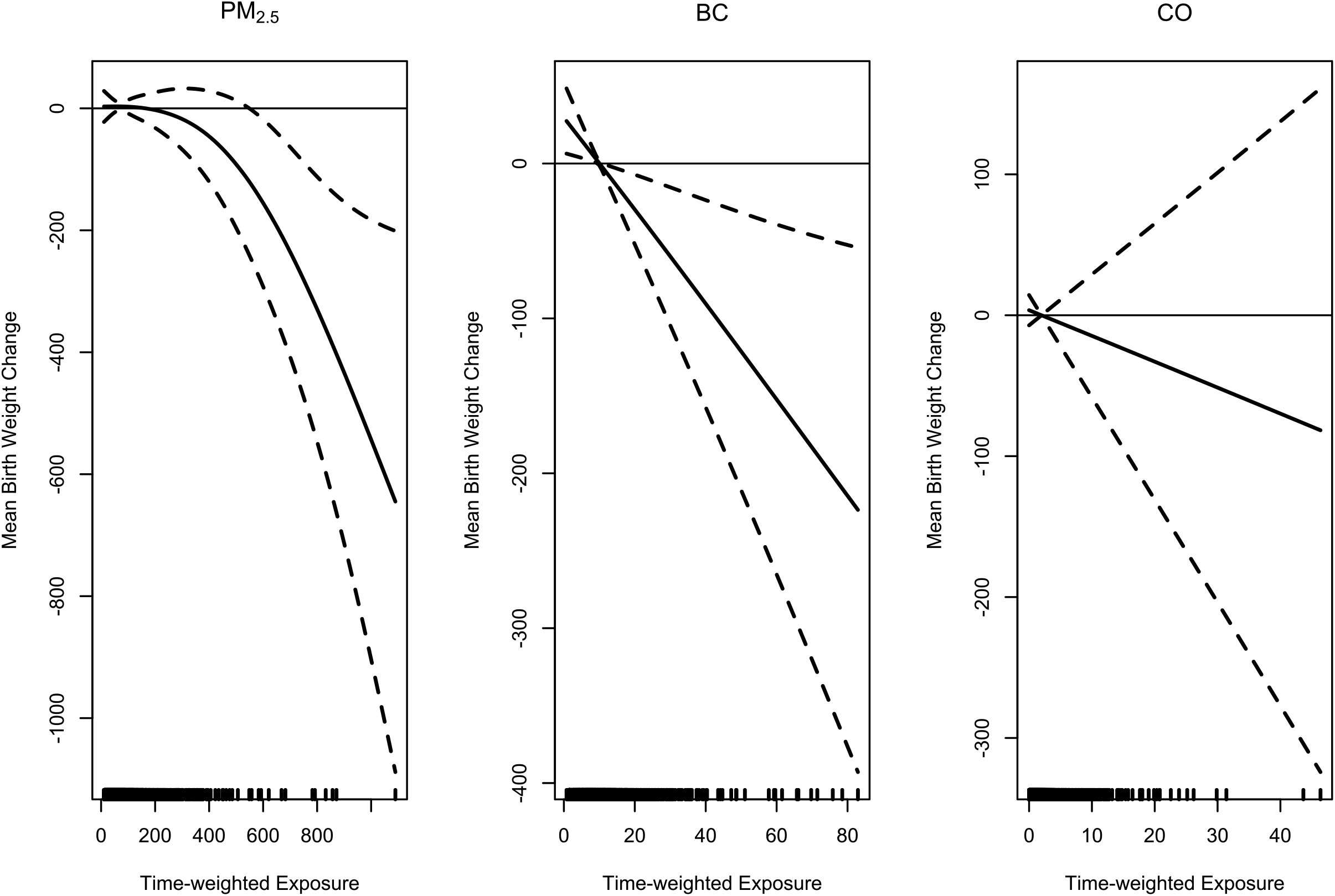
Exposure-response relationships between birthweight and prenatal PM_2·5_, BC, and CO personal exposures. Vertical dashes along the x-axis are observed measurements.

**Figure 3.**
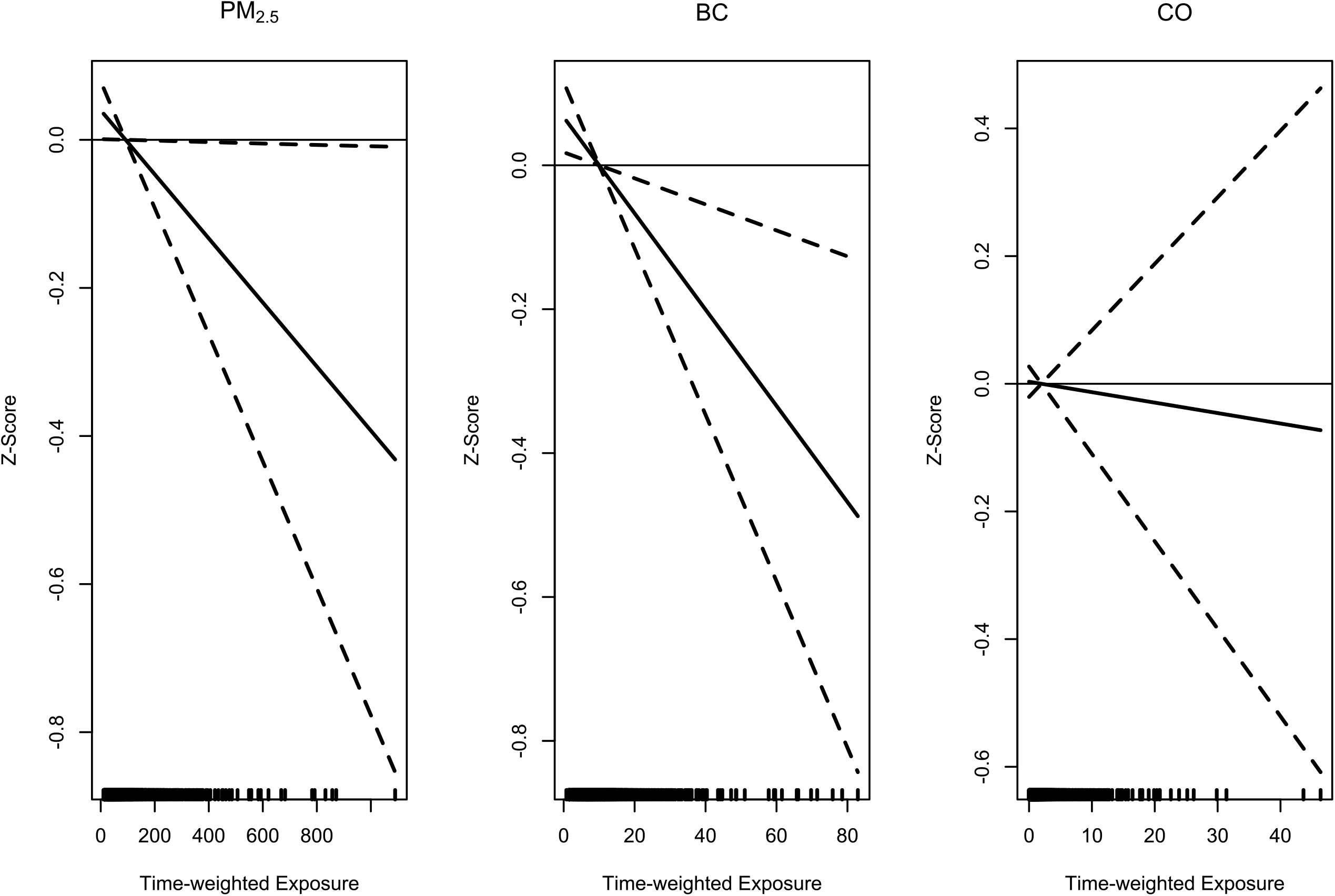
Exposure-response relationships between weight-for-gestational age z-scores and prenatal PM_2·5_, BC, and CO personal exposures. Vertical dashes along the x-axis are observed measurements.

## Discussion

The findings of this study suggest that reducing prenatal HAP exposure could yield modest potential benefits for birthweight that are not consistent across all pollutants. To our knowledge, ours is the first study reporting on E-R relationships between gestational BC exposures from HAP and birthweight. Notably, a 7·3 μg/m^3^ reduction in prenatal BC exposure was associated with an increase in birthweight of about 22 g, which could have positive implications for populations with a high prevalence of low birthweight.

Only three prior studies have published quantitative E-R results for birth outcomes in relation to HAP exposure, focusing on PM_2·5_ and/or CO. In a cohort of 239 pregnant women in Tanzania, there was a negative association between CO exposure and new-born birthweight, but results were not statistically significant.^29^ The Tanzania study also reported a 150 g (95% CI: −300 g, 0 g) reduction in birthweight per 23·0 μg/m^3^ increase in PM_2·5_. The second study, among 1285 women in the Tamil Nadu region of India, reported a 4 g (95% CI: 1·08 g, 6·76 g) decrease in birthweight and a 2% increase in the prevalence of LBW (95% CI: 0·05%, 4·1%) for each 10 μg/m^3^ increase in kitchen area PM_2·5_ measured during pregnancy.^10^ The third study, conducted as part of the GRAPHS trial in Ghana,^13^ observed effects of CO on birthweight, birth length, and gestational age that were modified by placental malarial status. Among infants from pregnancies without evidence of placental malaria, each 1 ppm increase in CO was associated with reduced birthweight (−53·4 g [95% CI: −84·8, −21·9 g]), birth length (−0·3 cm [95% CI: −0·6, −0·1 cm]), gestational age (−1·0 days [95% CI: −1·8, −0·2 days]), and weight-for-gestational age z-score (−0·08 [95% CI: −0·16, −0·01] standard deviations]). These associations were not observed in pregnancies with evidence of placental malaria. PM_2·5_ measurements were, however, limited in the GRAPHS trial and no association between PM_2·5_ exposure and birthweight was observed.

The negative associations between PM_2·5_ exposures and birthweight in our study are consistent with previous studies, but at the lower end of reported estimates. In contrast, the lack of an association between prenatal CO exposure and birthweight was unexpected. However, this is not entirely surprising as the correlations between PM_2·5_ and CO have not been uniform across HAP settings. A systematic review examining this relationship^30^ found inconsistent correlation with slightly stronger correlation among exclusive biomass users relative to mixed fuel users (*R*2 = 0·29 versus 0·18). The relatively modest correlations between either PM_2·5_ or BC and CO observed in our study may have been driven under our study conditions of exclusive biomass and LPG use.

Trials of cookstove interventions to improve birth outcomes have had mixed outcomes: an improved biomass cookstove in a cohort of 174 infants in Guatemala was associated with 89 g higher birth weight (95% CI: −27 g, 204 g) in adjusted analysis ^14^, and a clean-burning ethanol stove intervention in Nigeria was associated with 128 g higher birth weight (95% CI: 20 g, 236 g) among 258 infants in adjusted analysis.^12^ Meanwhile, neither an improved biomass nor an LPG stove improved birth outcomes in two linked trials covering almost 3000 individuals in southern Nepal.^15^ These trials have not reported quantitative E-R relations. In the GRAPHS trial, while there was a significant E-R relationship between CO exposures and birthweight,^13^ neither prenatally-introduced LPG nor improved biomass cookstoves improved birthweight. The investigators in all previous trials hypothesize that this is perhaps due to lower-than-expected exposure reductions in the intervention arm.

The HAP exposure levels associated with biomass use (such as at baseline and in the control arm) in our study are at the lower end of what has been reported in previous trials, with the possible exception of the GRAPHS trial. Based on pilot phase exposure reductions ^20,21^ and estimated supra-linear E-R relationships for HAP and birthweight,^31^ we hypothesised that the levels observed during pilot work implied that exposure reductions would occur on the steep part of the response curve for birthweight. Given the relative paucity of studies on quantitative E-R analyses for HAP based on personal exposures, it is quite possible that the shape of the E-R curve is different than what was previously estimated. Our study contributes important information regarding this relationship based on high quality personal HAP exposure and birthweight measurements from four diverse settings that can inform future development of pooled E-R coefficients spanning the range of experienced HAP exposures and may inform future E-R curves that integrate across air pollution sources.

Finally, we note that other unmeasured factors including placental malaria, water and sanitation, and nutritional deficiencies may have outweighed the effects of HAP on birthweight outcomes.

## Conclusions

In this study population drawn from diverse socio-demographic settings across four countries, exposure to HAP – particularly to BC and to a lesser extent to PM_2·5_ during pregnancy was associated with reduced birthweight and weight-for-gestational age z-scores. To our knowledge, ours is the first study reporting on E-R relationships between gestational BC exposures from HAP and birthweight. The association, while modest, provides strong support for continuing efforts to address HAP exposures alongside other drivers of impaired fetal growth in LMICs.

## Data Availability

All data produced in the present study are available upon reasonable request to the authors

http://sco.library.emory.edu/dataverse/

## Data Sharing

|  |  |
| --- | --- |
| Will individual participant data be available (including data dictionaries)? | Yes |
| What data in particular will be shared? | Individual participant data that underlie the results reported in this article, after de-identification (all results summarized in text, tables, figures, and appendices) |
| What other documents will be available? | Study protocol, statistical analysis plan, informed consent form, analytic code |
| When will data be available (start and end dates)? | Beginning 6 months following article publication |
| With whom? | Anyone who wishes to access the data |
| For what types of analyses? | Any purpose |
| By what mechanism will data be made available? | Data are available indefinitely at DataVerse (link to be included) |

## Acknowledgments

The HAPIN trial is funded by the U.S. National Institutes of Health (cooperative agreement 1UM1HL134590) in collaboration with the Bill & Melinda Gates Foundation (OPP1131279). The investigators would like to thank the members of the advisory committee - Patrick Brysse, Donna Spiegelman, and Joel Kaufman - for their valuable insight and guidance throughout the implementation of the trial. We also wish to acknowledge all research staff and study participants for their dedication to and participation in this important trial.

A multidisciplinary, independent Data and Safety Monitoring Board (DSMB) appointed by the National Heart, Lung, and Blood Institute (NHLBI) monitors the quality of the data and protects the safety of patients enrolled in the HAPIN trial. NHLBI DSMB: Nancy R Cook, Stephen Hecht, Catherine Karr (Chair), Joseph Millum, Nalini Sathiakumar, Paul K Whelton, Gail Weinmann and Thomas Croxton (Executive Secretaries). Program Coordination: Gail Rodgers, Bill & Melinda Gates Foundation; Claudia L Thompson, National Institute of Environmental Health Science; Mark J. Parascandola, National Cancer Institute; Marion Koso-Thomas, Eunice Kennedy Shriver National Institute of Child Health and Human Development; Joshua P Rosenthal, Fogarty International Center; Conception R Nierras, NIH Office of Strategic Coordination Common Fund; Katherine Kavounis, Dong-Yun Kim, Antonello Punturieri, and Barry S Schmetter, NHLBI. The findings and conclusions in this report are those of the authors and do not necessarily represent the official position of the US National Institutes of Health or Department of Health and Human Services.

