## Supplementary Material for "Exposure–response relationships for personal exposure to fine particulate matter (PM_2·5_), carbon monoxide, and black carbon and birthweight: Results from the multi-country Household Air Pollution Intervention Network (HAPIN) trial"


Table S1: Change in birthweight for by quartiles of exposure 2
Table S2: Change in z-scores by quartiles of exposure 2
Table S3. AICs for different models for birthweight and z-score, in relation to $PM_{2\cdot5}$ and BC 3
Table S4. Change in outcome by increase in pollutant IQR for full term births 3
Table S5. Change in outcome by increase in pollutant IQR by infant sex 3
Table S6: Birthweights and pollutant exposures by country and overall 4
Table S7. Linear models for birthweight and three pollutants in each country 5
Table S8. Linear models for weight-for gestational age z-scores and three pollutants in each country 5
Table S9. Exposure-response coefficient using exposure as measured in early, middle, and late gestation 6
Figure S1. CONSORT diagram 7
Figure S2. Graphs of linear, quadratic, 2-piece linear and cubic spline models for $PM_{2\cdot5}$ and birthweight 8
Figure S3. Directed acyclic graph (DAG) to guide the selection of confounders for the association between household air pollution exposure and birthweight. 8

**Table S1: Change in birthweight for by quartiles of exposure***

| Pollutant | Model | Estimate (95% CI) | p-value | AIC |
|---|---|---|---|---|
| $PM_{2·5}$ | Quartile 1 | *reference* | | 40218 |
| | Quartile 2 | -24·9 (-68·9, 19·2) | 0·27 | |
| | Quartile 3 | -5·2 (-50·1, 39·7) | 0·82 | |
| | Quartile 4** | -24·5 (-70·4, 21·4) | 0·30 | |
| BC | Quartile 1 | *reference* | | 37885 |
| | Quartile 2 | -29·9 (-75·3, 15·5) | 0·20 | |
| | Quartile 3 | -18·3 (-64·9, 28·3) | 0·44 | |
| | Quartile 4** | -39·2 (-86·1, 7·7) | 0·10 | |
| CO | Quartile 1 | *reference* | | |
| | Quartile 2 | 44·0 (1·7, 86·2) | 0·04 | |
| | Quartile 3 | 47·4 (4·9, 89·9) | 0·03 | |
| | Quartile 4** | 16·1 (-27·3, 59·5) | 0·47 | 41015 |

*All models adjusted for mother's education, baseline BMI, nulliparity, diet diversity, food insecurity score, second-hand smoke, baseline hemoglobin, age, as well as infant sex, and 10 randomization strata.
**Cutpoints for quartiles for $PM_{2·5}$: 40·8, 69·5, and 115·3cutpoints for quartiles for BC: 5·4, 8·6, and 12·7, cutpoints for quartiles for CO: 0·55, 1·13, and 2·23.

**Table S2: Change in z-scores by quartiles of exposure***

| Pollutant | Model | Estimate (95% CI) | p-value | AIC |
|---|---|---|---|---|
| $PM_{2·5}$ | Quartile 1 | *reference* | | 7024 |
| | Quartile 2 | -0·094 (-0·191, 0·002) | 0·06 | |
| | Quartile 3 | -0·045 (-0·144, 0·054) | 0·37 | |
| | Quartile 4** | -0·102 (-0·202, 0·001) | 0·05 | |
| BC | Quartile 1 | *reference* | | 6595 |
| | Quartile 2 | -0·063 (-0·163, 0·037) | 0·21 | |
| | Quartile 3 | -0·076 (-0·179, 0·023) | 0·14 | |
| | Quartile 4** | -0·137 (-0·240, -0·034) | 0·01 | |
| CO | Quartile 1 | *reference* | | 7212 |
| | Quartile 2 | 0·013 (0·038, 0·225) | 0·01 | |
| | Quartile 3 | 0·097 (0·003, 0·192) | 0·04 | |
| | Quartile 4** | 0·060 (-0·037, 0·156) | 0·23 | |

*All models adjusted for mother's education, baseline BMI, nulliparity, diet diversity, food insecurity score, second-hand smoke, baseline hemoglobin, age, infant sex, and 10 randomization strata.
**Cutpoints for quartiles for PM2·5: 40·8, 69·5, and 115·3cutpoints for quartiles for BC: 5·4, 8·6, and 12·7, cutpoints for quartiles for CO: 0·55, 1·13, and 2·23.

**Table S3. AICs for different models for birthweight and z-score, in relation to $PM_{2\cdot5}$ and BC**

| Outcome | Exposure | Model | AIC |
|---|---|---|---|
| birthweight | $PM_{2\cdot5}$ | linear | 40211·7 |
| | | Log linear | 40215·0 |
| | | *quadratic* | 40206·2 |
| | | 2 piece linear | 40207·4 |
| | | Cubic spline | 40212·4 |
| birthweight | BC | *linear* | 37876·6 |
| | | Log linear | 37880·8 |
| | | quadratic | 37878·2 |
| | | 2 piece linear | 37877·5 |
| | | Cubic spline | 37878·2 |
| z-score | $PM_{2\cdot5}$ | *linear* | 7021·6 |
| | | Log linear | 7023·2 |
| | | quadratic | 7023·5 |
| | | 2 piece linear | 7022·9 |
| | | Cubic spline | 7023·6 |
| z-score | BC | *linear* | 6590·7 |
| | | Log linear | 6592·8 |
| | | quadratic | 6592·5 |
| | | 2 piece linear | 6592·3 |
| | | Cubic spline | 6592·5 |

**Table S4. Change in outcome by increase in pollutant IQR for full term births**

| Outcome | Pollutant and model | Change in outcome (g) | LCL change | UCL change |
|---|---|---|---|---|
| birthweight | $PM_{2\cdot5}$ linear | -13·315 | -27·017 | 0·395 |
| | BC linear | -19·701 | -34·757 | -4·645 |
| | CO linear | -3·183 | -11·570 | 5·204 |
| z-score | $PM_{2\cdot5}$ linear | -0·045 | -0·075 | -0·015 |
| | BC linear | -0·054 | -0·089 | -0·018 |
| | CO linear | -0·006 | -0·025 | 0·014 |

**Table S5. Change in outcome by increase in pollutant IQR by infant sex**

| Outcome | Pollutant and model | Change in outcome (g) | LCL change | UCL change |
|---|---|---|---|---|
| birthweight (male) | $PM_{2\cdot5}$ linear | -2·776 | -23·415 | 17·863 |
| | BC linear | -7·871 | -31·385 | 15·643 |
| | CO linear | -6·144 | -19·729 | 7·441 |
| birthweight (female) | $PM_{2\cdot5}$ linear | -26·674 | -46·009 | -7·339 |
| | BC linear | -34·745 | -56·388 | -13·103 |
| | CO linear | -0·081 | -12·063 | 11·901 |
| z-score (male) | $PM_{2\cdot5}$ linear | -0·032 | -0·076 | 0·012 |
| | BC linear | -0·040 | -0·090 | 0·010 |
| | CO linear | -0·010 | -0·039 | 0·020 |
| z-score (female) | $PM_{2\cdot5}$ linear | -0·031 | -0·075 | 0·012 |
| | BC linear | -0·059 | -0·108 | -0·010 |
| | CO linear | 0·002 | -0·026 | 0·029 |

**Table S6: Birthweights and pollutant exposures by country and overall**

| | Birthweight | | | $PM_{2\cdot5}$ | | | | BC | | | | CO | | | |
|---|---|---|---|---|---|---|---|---|---|---|---|---|---|---|---|
| | **Mean** | **SD** | **N** | **Mean** | **SD** | **Median** | **N** | **Mean** | **SD** | **Median** | **N** | **Mean** | **SD** | **Median** | **N** |
| India | 2591 | 394 | 773 | 94·29 | 90·90 | 69·32 | 710 | 10·44 | 8·49 | 8·24 | 698 | 1·49 | 2·21 | 0·87 | 735 |
| Rwanda | 3022 | 439 | 749 | 93·50 | 72·45 | 75·42 | 695 | 10·42 | 6·38 | 9·19 | 618 | 1·94 | 2·66 | 1·04 | 710 |
| Guatemala | 2861 | 428 | 750 | 112·88 | 83·89 | 88·62 | 703 | 10·90 | 6·40 | 9·94 | 677 | 1·56 | 1·64 | 1·08 | 727 |
| Peru | 3180 | 410 | 730 | 64·37 | 79·85 | 38·99 | 609 | 7·96 | 7·71 | 5·75 | 567 | 3·09 | 4·41 | 1·76 | 600 |
| Study-wide | 2909 | 471 | 2560 | 92·19 | 83·85 | 69·45 | 2717 | 10·01 | 7·39 | 8·57 | 2560 | 1·97 | 2·90 | 1·13 | 2772 |

**Table S7. Linear models for birthweight and three pollutants in each country**

| Pollutant and model | Country | Exposure-response coefficient | p-value |
|---|---|---|---|
| $PM_{2·5}$ linear model | Guatemala | -0·11 | 0·55 |
| | India | -0·09 | 0·57 |
| | Peru | -0·16 | 0·44 |
| | Rwanda | -0·56 | 0·01 |
| | Combined | -0·20 | 0·04 |
| BC linear model | Guatemala | -4·5 | 0·07 |
| | India | -2·6 | 0·13 |
| | Peru | -0·9 | 0·69 |
| | Rwanda | -3·5 | 0·22 |
| | Combined | -3·0 | 0·01 |
| CO linear model | Guatemala | -10·6 | 0·26 |
| | India | 3·1 | 0·61 |
| | Peru | -5·6 | 0·13 |
| | Rwanda | 10·9 | 0·24 |
| | Combined | -1·8 | 0·49 |

**Table S8. Linear models for weight-for-gestational age z-scores and three pollutants in each country**

| Pollutant and model | Country | Exposure-response coefficient | p-value |
|---|---|---|---|
| $PM_{2·5}$ linear model | Guatemala | -0·0005 | 0·29 |
| | India | -0·0004 | 0·25 |
| | Peru | -0·0002 | 0·61 |
| | Rwanda | -0·0006 | 0·23 |
| | Combined | -0·0004 | 0·04 |
| BC linear model | Guatemala | -0·125 | 0·03 |
| | India | -0·006 | 0·09 |
| | Peru | -0·001 | 0·76 |
| | Rwanda | -0·005 | 0·37 |
| | Combined | -0·007 | 0·01 |
| CO linear model | Guatemala | -0·006 | 0·79 |
| | India | 0·011 | 0·40 |
| | Peru | -0·010 | 0·23 |
| | Rwanda | 0·009 | 0·49 |
| | Combined | -0·002 | 0·78 |

**Table S9. Exposure-response coefficient using exposure as measured in early, middle, and late gestation**

| | Coefficient | t-statistic | p-value |
|---|---|---|---|
| **Birthweight** | | | |
| BC overall avg | -2·9996 | 7·42 | 0·0065 |
| BC BL* | -0·856 | 1·06 | 0·3044 |
| BC P1* | -1·8725 | 3·38 | 0·0661 |
| BC P2* | -1·8086 | 3·62 | 0·0571 |
| $PM_{2·5}$ overall avg | -0·1983 | 4·3 | 0·0381 |
| $PM_{2·5}$ BL | -0·07 | 1·15 | 0·2843 |
| $PM_{2·5}$ P1 | -0·0033 | 0 | 0·9728 |
| $PM_{2·5}$ P2 | -0·0645 | 0·51 | 0·4761 |
| CO overall avg | -1·8209 | 0·46 | 0·4999 |
| CO BL | 2·3535 | 1·59 | 0·2076 |
| CO P1 | 2·1364 | 0·61 | 0·4357 |
| CO P2 | -3·7919 | 2·13 | 0·1448 |
| **Weight-for-gestational age z-scores** | | | |
| BC overall avg | -0·0067 | 7·62 | 0·0058 |
| BC BL* | -0·0028 | 2·41 | 0·1202 |
| BC P1* | -0·004 | 3·09 | 0·0786 |
| BC P2* | -0·0043 | 4·17 | 0·0411 |
| $PM_{2·5}$ overall avg | -0·0004 | 4·32 | 0·0377 |
| $PM_{2·5}$ BL | -0·0002 | 2·61 | 0·1063 |
| $PM_{2·5}$ P1 | -0·0001 | 0·19 | 0·6659 |
| $PM_{2·5}$ P2 | -0·0002 | 0·88 | 0·3475 |
| CO overall avg | -0·0017 | 0·08 | 0·7787 |
| CO BL | 0·0064 | 2·38 | 0·1226 |
| CO P1 | 0·0043 | 0·5 | 0·4794 |
| CO P2 | -0·0073 | 1·58 | 0·2089 |

*BC = black carbon, CO = carbon monoxide
**BL = baseline measurement, P1 and P2 are 2nd and 3rd measurements

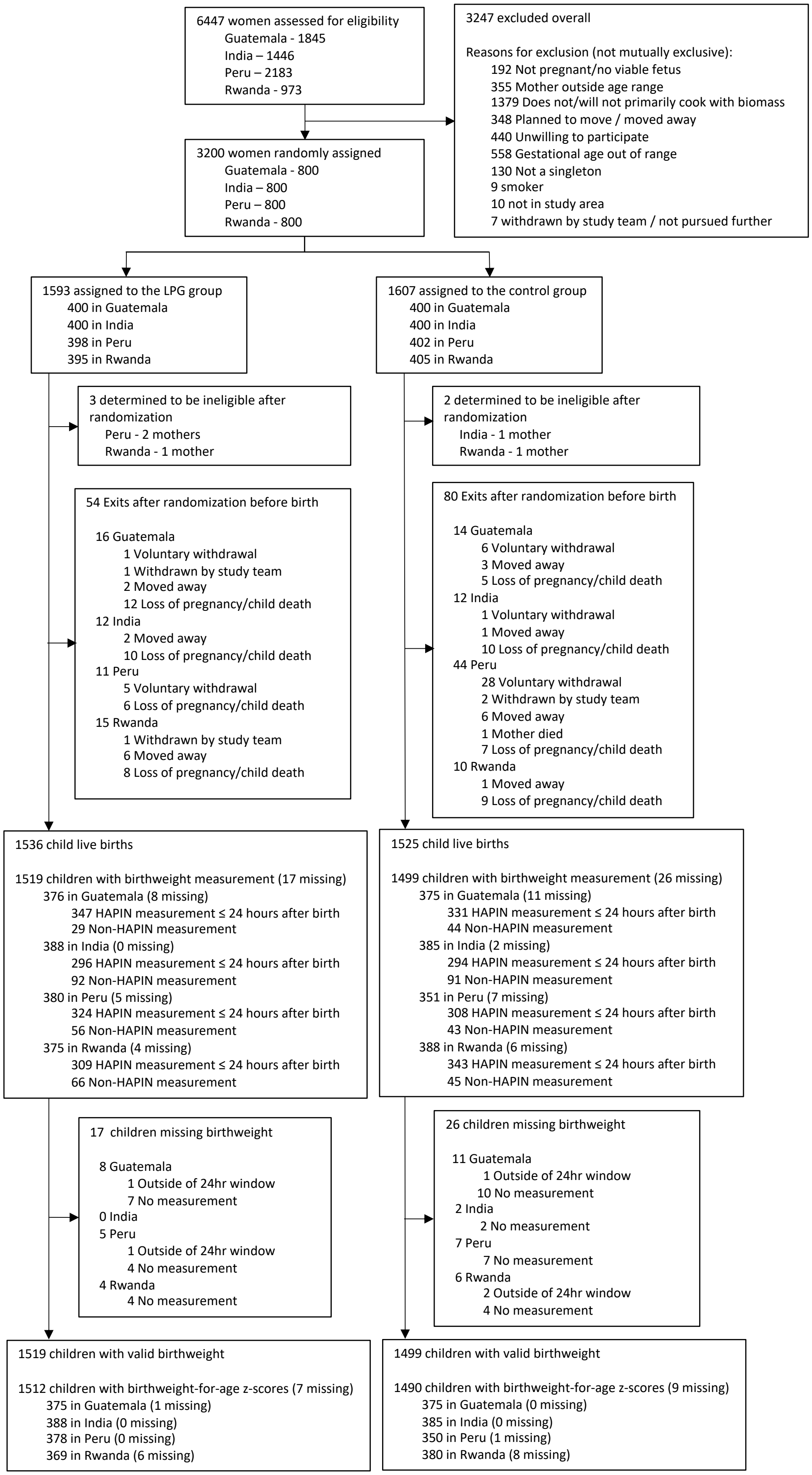


**Figure S1. CONSORT diagram**

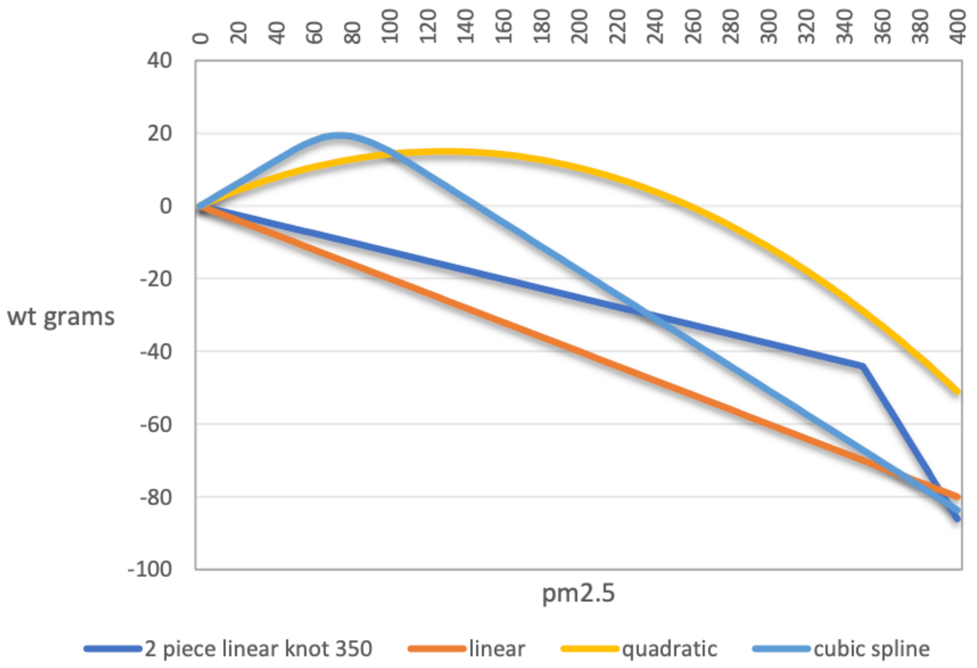


**Figure S2. Linear, quadratic, 2-piece linear and cubic spline models for $PM_{2.5}$ and birthweight** Linear model AIC 40212; 2-piece spline model knot 350, AIC 40207; Quadratic AIC 40206. Cubic spline model AIC 40212.

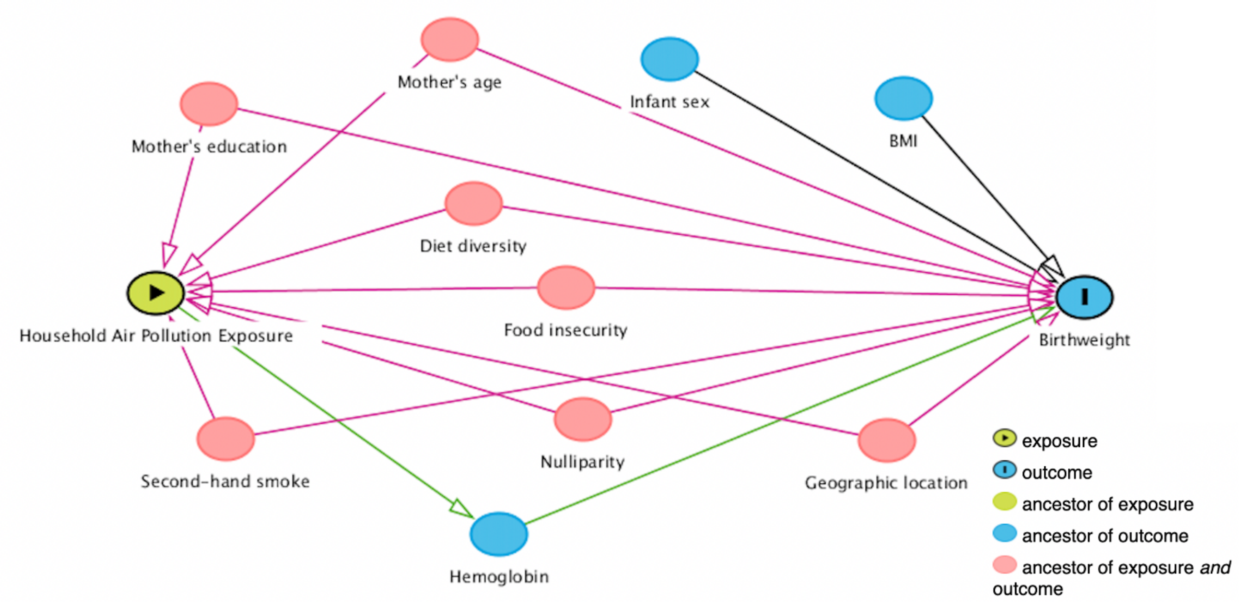


**Figure S3.** Directed acyclic graph (DAG) of the a priori covariates adjusted in the exposure-response models for the associations between household air pollution exposure and birthweight. Causal path (green line); Biasing path (red line).
